# Research on the relationship between postoperative D-dimer clearance and trauma infection complications in patients with different degrees of fracture

**DOI:** 10.1101/2020.02.21.20024737

**Authors:** Hai-Mei Ma, Yong-wei Pan, Lianxu Chen

## Abstract

**Background:** Occurrence of thrombotic disease after orthopedic surgery has always been an important issue affecting the prognosis of patients. In this paper, retrospective analysis was used to analyze the D-dimer clearance rate after different degrees of fractures, suggesting that the D-dimer clearance rate can be used as an index to evaluate thrombotic diseases after orthopedic surgery.

**Material/Methods:** Seventy-five patients with orthopedic surgery were randomly selected from March to June 2017. According to the degrees of fractures and complications, they were divided into SF(single fracture), MF(multiple fracture), FCI(Fracture combined with infection)three groups, 25 in each group.D-dimer results of each case on 1 day, 2 days, 3 days, 4 days, 5 days, and 7 days after orthopedic surgery were recorded and counted.The slope of correlation equation of D-dimer value according to time is the D-dimer clearance rate.

**Results:** The D-dimer clearance rate in the SF□JMF □FCI group was −0.0490, −0.0502 and −0.0692□The P value is respectively 0.0049, 0.0061 and 0.0163, the difference is significant.

**Conclusions:** After traumatic fracture surgery, whether D-dimer clears at a normal rate is closely related to its outcome. Decreasing D-dimer clearance is related to postoperative infections and thrombotic diseases. D-dimer clearance can be used as an important parameter and observation index for judging the clinical outcome of patients with complications such as severe trauma and infection. At the same time, we can further study the cut-off value of D-dimer clearance.

## Background

With the improvement of treatment and rescue, the life of patients with traumatic fractures is greatly guaranteed. However, thrombotic diseases such as PE (pulmonary embolism), DVT (deep vein thrombosis), etc. after orthopedic surgery are still important issues affecting the prognosis of patients. Clinicians place great emphasis on postoperative complications and related laboratory tests.

Many laboratory indicators about coagulation system and laboratory methods for the diagnosis of venous thrombosis have been discovered with the development of monoclonal antibody technology, of which, D-dimer is the most widely used and sensitive parameter[1].D-dimer is a specific peptide segment in which plasmin degrades cross-linked fibrin[2].Its increased content indicates the secondary increased fibrinolytic activity caused by hypercoagulability in the body, suggesting active thrombosis, which is used as one of the indicators for screening perioperative thrombus[3]. In view of this,we have introduced a new indicator of D-dimer clearance in this article, we analyzed postoperative D-dimer clearance in patients with varying degrees of fracture and explored the association between D-dimer clearance rate and postoperative patient complications.

## Material and Methods

### Study design

This study is a retrospective analysis.Statistical methods were used to estimate the amount of sample required for this observational study. Seventy-five patients with orthopedic surgery were randomly selected from March to June 2017. According to the degree of fracture injury and complications, they were divided into three groups, 25 in each group.

In the SF(single fracture) group,including 13 males and 12 females, aged 15-68 years (mean age: 30.2 ±11.2 years);in the MF(multiple fracture) group (number of fractures≥2),including 15 males and 10 females, aged 24-79 years (mean age:45.3 ±13.1); in the FCI(Fracture combined with infection) group,including 17 males and 8 females, aged 35-70 years (mean age:55.3 ±10.3 years).Each case D-dimer results of 1 day, 2 days, 3 days, 4 days, 5 days, and 7 days after orthopedic surgery were recorded and counted.

Cases with incomplete D-dimer results were excluded. All patients had no history of abnormal blood coagulation and abnormal fibrinolytic activity. Whole blood and plasma were not infused during and after surgery. All patients underwent continuous epidural anesthesia. No other drugs were added during the operation. Only saline and colloidal equilibrium solution were entered.

### Supplemental Methods

The study was approved by local institutional ethic committees, registered under the National Institute for clinical trial (http://218.240.145.213:9000/CTMDS/apps/pub/public.jsp), ethical review approval numberof this study is:16152-110.All participants provided written informed consent.

### Specimen collection

Peripheral venous blood was drawn on 1d, 2d, 3d, 4d, 5d and 7d after operation respectively, and anticoagulated with 0.108 mol/L sodium citrate at a ratio of venous blood to anticoagulant of 9:1, and then mixed well immediately, centrifuged for 10 min at 3000 r/min to separate platelet-poor plasma for D-dimer detection.

### Parameters to be determined

D-dimer was tested by an Innovance D-dimer test kit provided by Semins, USA. According to the method for “excluding thrombotic diseases” recommended by Clinical & Laboratory Standards Institute (CLSI) H59-P.

Instrument: Sysmex CA-7000 automatic coagulation analyzer,Made in Japan.

Method: D-dimer in plasma was determined by immunoturbidimetric assay.

D-dimer Clearance rate :Obtained from the slope of D-dimer with time-dependent equation.

### Evaluation Criteria

When the D-dimer level was >0.5mg/l FEU, it was a positive result (<0.5mg/l FEU is the normal reference range of Innovance D-dimer level).

Classification of fractures: Multiple fractures meant that fractures occurred in two or more sites. More fractures in the same site, such as multiple rib fractures, pubic fractures, sciatic fractures, or injuries caused by the same external force mechanism, such as MaisOn-neuve fracture, Monteggia fracture and Galeazzi fracture, were calculated by a single injury.

Judgment of co-infection: The pathogenic bacteria were isolated and identified according to definite clinical diagnosis.

The outcome of postoperative traumatic fractures was considered in terms of hospitalization time and postoperative thrombotic diseases and infections.

D-dimer Clearance rate is expressed as the slope of the correlation curve of the D-dimer level at different times.

### Statistical Processing

Statistical analysis was performed using SPSS 23 software.

The D-dimer mean values of different groups and different time were compared by t test. The clearance rates of different groups of D-dimer were compared by chi-square test.

## Results

### General information of cases selected

The number, age and sex composition of cases were shown in the Table 1. The postoperative outcome was evaluated by the average length of stay. Patients in the infection group were transfered to the infection ward or ICU for observation and treatments after definite diagnosis.

**Table 1.** General information of observation groups.

|  | SF group | MF group | FCI group | Total |
| --- | --- | --- | --- | --- |
| Number of cases | 25 | 25 | 25 | 75 |
| Age,y | 30.2 ±11.2 | 45.3 ±13.1 | 55.3 ±10.3 |  |
| Gender |  |  |  |  |
| Male | 13 | 15 | 17 | 45 |
| Female | 12 | 10 | 8 | 30 |
| hospital stay(Days) | <9 | >14 | >28 |  |

SF=Single fracture (number of fractures <2); MF=Multiple fracture (number of fractures ≥2); FCI=Fracture combined with infection (isolation of pathogenic bacteria after definite clinical diagnosis)

### The mean level of D-dimer at different time after orthopedic surgery

Mean level of D-dimer at different time after orthopedic surgery was shown in Table 2.In SF group, the levels of D-dimer 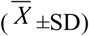 (mg/l FEU) on 1 day, 2 days, 3 days, 4 days, 5 days, 7 days after surgery were 8.36±0.41, 5.73±0.32, 4.2±0.23, 2.78±0.13, 1.83±0.09, 1.15±0.06; In the MF group, the levels of D-dimer 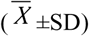 (mg/l FEU) on 1 day, 2 days, 3 days, 4 days, 5 days, 7 days after surgery were 15.07±0.91, 12.03±0.72, 10.4±0.56, 9.2±0.47, 8.4±0.39, 7.44±0.39; and in the FCI group, the levels of D-dimer 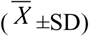 (mg/l FEU)on 1 day, 2 days, 3 days, 4 days, 5 days, 7 days after surgery were 23.11±1.21, 17.76±0.92, 14.47±0.77, 15.63±0.21, 14.24±0.79, 11.38±0.68.The change curve of mean level of D-dimer at different time after orthopedic surgery in each group was shown in Figure 1,Innovance D-dimer<0.5mg/l FEU was used as the upper limit of the normal reference range. The average values of D-dimer in SF, MF and FCI groups were positive results, indicated by * in the table 2.

**Table 2.** Mean level of D-dimer at different time after orthopedic surgery (mg/l FEU)

| D-dimer( $\bar{X} \pm SD$) | SF group | MF group | FCI group |
| --- | --- | --- | --- |
| Days(Hours) postoperation |  |  |  |
| 1(24) | 8.36±0.41* | 15.07±0.91* | 23.11±1.21* |
| 2(48) | 5.73±0.32* | 12.03±0.72* | 17.76±0.92* |
| 3(72) | 4.2±0.23* | 10.4±0.56* | 14.47±0.77* |
| 4(96) | 2.78±0.13* | 9.2±0.47* | 15.63±0.21* |
| 5(120) | 1.83±0.09* | 8.4±0.39* | 14.24±0.79* |
| 7(168) | 1.15±0.06* | 7.44±0.39* | 11.38±0.68* |
| T-test ( P ) | SF-MF 0.000<sup>#</sup> | MF-FCI 0.004<sup>#</sup> | SF-FCI 0.002<sup>#</sup> |

SF=Single fracture (number of fractures <2); MF=Multiple fracture (number of fractures ≥2); FI=Fracture with infection (isolation of pathogenic bacteria after definite clinical diagnosis); "\*" positive results; "<sup>#</sup>" Significant difference( $P < 0.01$ )

**Figure 1.**
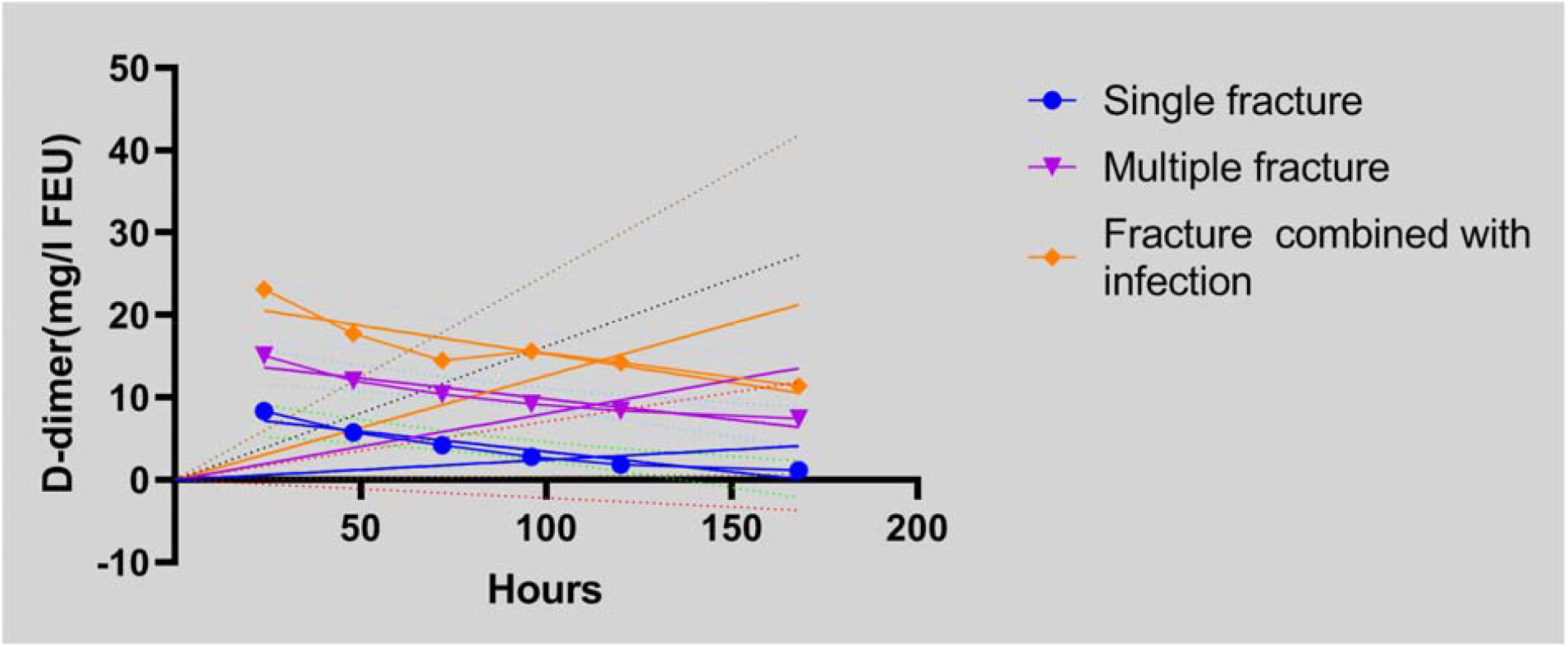
Change curves of mean levels of D-dimer at different time after orthopedic surgery.

### D-dimer clearance rates after orthopedic surgery

The changing rate of D-dimer at different times after orthopedic surgery means D-dimer clearance rate.We obtain the correlation equation between D-dimer value and time according to the D-dimer values of SF, MF and FCI groups.The slope of this equation means the changing rate of D-dimer, which is the clearance rate of D-dimer.The correlation curves and related equations of D-dimer in SF, MF and FCI groups were shown in Figure 2-4. D-dimer clearance rate was shown in Table 3.

**Table 3.** D-dimer clearance rates of observation groups after orthopedic surgery.

|  | SF group | MF group | FCI group |
| --- | --- | --- | --- |
| DCR(Slope) | -0.049 | -0.0502 | -0.0692 |
| Equation | $Y = -0.0490 * X + 8.321$ | $Y = -0.0502 * X + 14.840$ | $Y = -0.0692 * X + 22.187$ |
| Y-intercept | 8.321 | 14.840 | 22.187 |
| X-intercept | 169.8 | 295.7 | 320.7 |
| 1/slope | -20.41 | -19.93 | -14.45 |
| R square | 0.8877 | 0.8754 | 0.799 |
| F | 31.61 | 28.12 | 15.9 |
| P value | SF-FCI 0.0049<sup>#</sup> | MF-FCI 0.0061<sup>#</sup> | SF-MF 0.0163<sup>Δ</sup> |

DCR=D-dimer clearance rates ; “<sup>#</sup>” Significant difference ( $P<0.01$ ) ; “<sup>Δ</sup>” no Significant difference ( $P>0.01$ )

**Figure 2.**
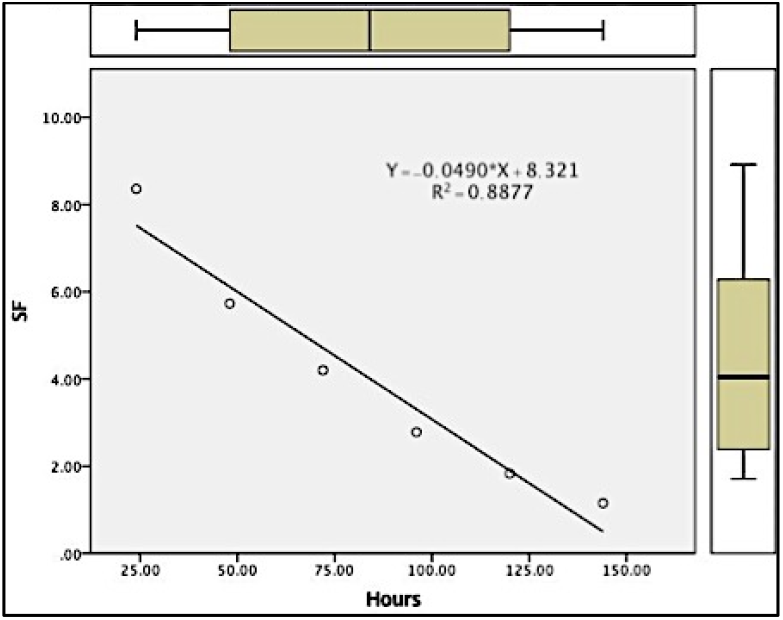
D-dimer clearance rates of SF group.

**Figure3.**
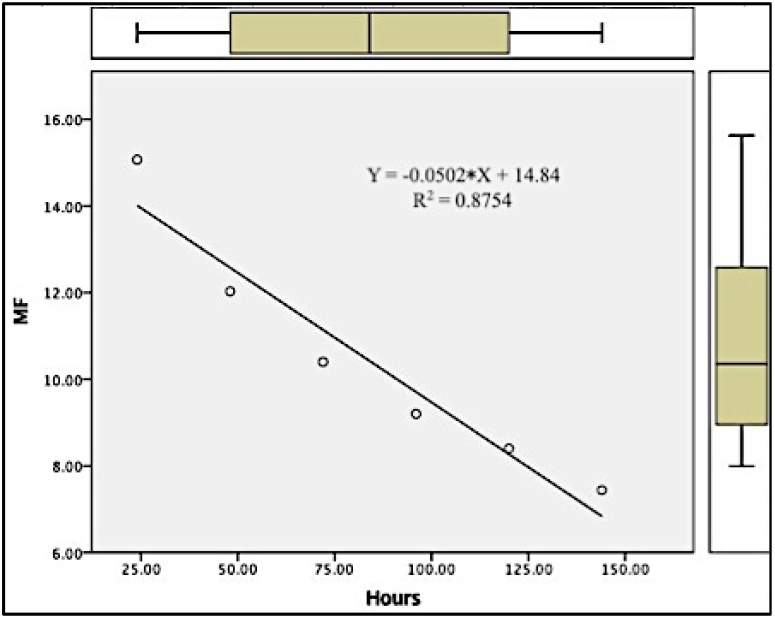
D-dimer clearance rates of MF group.

**Figure4.**
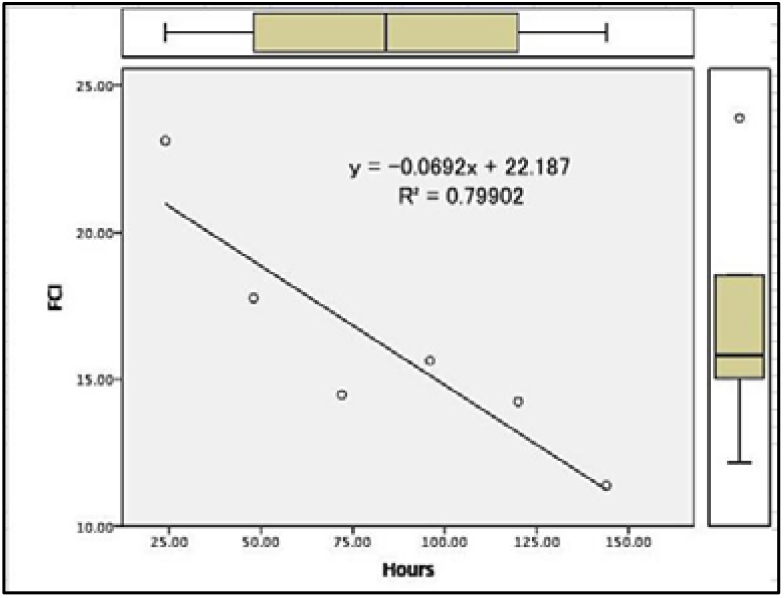
D-dime clearance rates of FCI group.

The clearance rate of D-dimer at different times in the SF,MF,FI group was −0.049,-0.0502 and −0.0692,The *P* value is respectively 0.0049,0.0061 and 0.0163.There were significant differences in the clearance rates of D-dimer between the SF group and the FCI group (*P*<0.01), and between the MF group and FCI group (*P*<0.01), The clearance rate of D-dimer in the SF group and MF group was higher than that of the FCI group. There was no significant difference in the clearance rates of D-dimer between the SF group and the MF group (P>0.01).

## Discussion

The dynamic balance of the coagulation and anticoagulation systems is the basis for maintaining the normal state of blood in the blood vessels. The fibrinolytic system, as one of the anticoagulant pathways, consists of four main components: Plasminogen, plasminogen activator, plasmin and plasmin inhibitor. When the blood clotting activity is increased and the fibrin clot is formed, plasminogen is activated and converted to plasmin and the fibrin starts lysis under the action of plasminogen activator such as t-PA (tissue-type plasminogen activator). Cross-linked fibrin produces a most stable specific peptide D-dimer under the action of plasmin, with stronger antigenicity[4]. The elevated D-dimer plasma levels indicate the presence of fibrin-mediated secondary fibrinolysis, and the increased local fibrinolysis activity for cross-linked fibrin.At present, based on the mechanism of D-dimer formation, the detection of D-dimer level in plasma has become a direct and practical method for determining the risk of thrombosis [5], which can facilitate the monitoring of thrombosis.

Apparently elevated D-dimer level occurred after orthopedic surgery[6]. Owings[7], Chen JP[8], Chen Liu[9] et al. found that most patients had an elevated D-dimer level in plasma after orthopedic.Oshima K et al. applied the D-dimer level to the assessment of the severity of trauma and postoperative mortality and found that the D-dimer and FDP levels were significantly increased in the dead cases after trauma[10].The post-traumatic hemorrhage and elevated fibrinolytic activity were closely related to the occurrence of DIC (disseminated intravascular coagulation) [11]. Patients with pyemia have a significantly abnormal coagulation function compared with trauma patients[12]. In previous studies, we also found that D-dimer levels are closely related to the severity of trauma fractures[13].

In this paper, we investigated the postoperative dynamic changes of D-dimer using cases of orthopedic surgery, to obtain the rule of D-dimer changes (the D-dimer clearance rate) after the trauma fracture surgery and study the relationship between the D-dimer clearance rate and the occurrence of postoperative thrombotic diseases, postoperative outcome and complications. The D-dimer values at different time points in the SF, MF, and FCI groups were higher than the upper limit of the normal range (positive results) and the difference was extremely significant (p<0.01), which means that D-dimer increased after trauma fracture. It is closely related to the degree of fracture trauma and postoperative outcome, which is consistent with previous conclusions.

We first proposed the concept of D-dimer clearance rate. 450 D-dimer results of 75 cases were summarized and statistical analyzed.It showed that the D-dimer clearance rate in single fracture group and multiple fracture group was significantly difference to the fracture-infected group, which was higher than that in the fracture-infected group, indicating that new D-dimer was produced in the body, and the patient was in a state of excessive fibrinolysis, which was associated with the intravascular coagulation (DIC), multiple organ failure syndrome (MOFS) and acute respiratory distress syndrome (ARDS)[14].

D-dimer will generally increase after fracture trauma^[7]-[9]^, D-dimer positive increase has no specific diagnostic significance for thrombotic diseases after fracture trauma, so this paper starts from the postoperative D-dimer clearance rate to explore its thrombotic disease diagnostic value of infection complications. In order to get the closest D-dimer change value, we retrospectively analyzed the D-dimer at 1d-7d six postoperative time, and the correlation equation R square was related, but it was not ideal. In order to get a more realistic correlation equation R square, the observation time point should be more, which has more clinical application value.

In clinical practice, not every patient after trauma can dayly dynamically observe D-dimer changes. Therefore, the rate of D-dimer clearance is more significant for judging clinical outcomes in patients with complications such as severe trauma and infection. The patient’s outcome can be judged by changes in daily D-dimer results.

## Conclusion

After traumatic fracture surgery, whether D-dimer clears at a normal rate is closely related to its outcome. Decreasing D-dimer clearance is related to postoperative infections and thrombotic diseases. D-dimer clearance can be used as an important parameter and observation index for judging the clinical outcome of patients with complications such as severe trauma and infection. At the same time, we can further study the cut-off value of D-dimer clearance.

## Funding

Capital Characteristic Clinical Application Research Fund(CN)

Award Number:12017B3003

## Data Availability

All relevant data are within the paper and its Supporting Information files.

## Acknowledgement

Thanks to Director Zhang Xiaofei and Chen Peng for helping with statistical processing.

## Conflict of interest

None.

## Notes

### Competing Interest Statement

The authors have declared no competing interest.

### Clinical Trial

This study was registered with a clinical institution and received an ethical review. Ethical certification documents are attached to the supplementary documents.

## References

1. Kabrhel C,Mark Courtney D,Camargo CA Jr,Plewa MC,Nordenholz KE,Moore CL,et al.Factors associated with positive D-dimer results in patients evaluated for pulmonary embolism.Acad Emerg Med 2010;17:589–597.

2. Kelly J,Rudd A,Lewis RR,Hunt BJ.Plasma D-dimers in the diagnosis of venous thromboembolism.Arch-Inter-Med,2002. 747∼756.

3. Heim SW,Schectman JM,Sindaty MS,Philbrick JT.D-dimer testing for deep venous thrombosis:a meta analysis.Clin Chem 2004;50:1136–1147.

4. Kabrhel C,Mark Courtney D,Camargo CA Jr,Plewa MC,Nordenholz KE,Moore CL,et al.Factors associated with positive D-dimer results in patients evaluated for pulmonary embolism.Acad Emerg Med 2010,17:589–597.

5. Komberrg A,Carles WF,Victor JM.Plasma rosslinked fibrin polymers:Quantitation based on tissue plasminogen activator conversion to D-dimer and measurement in normal and patients with acute thrombotic disorders.Blood,1992, p709∼717.

6. Kelly J,Rudd A,Lewis RR,Hunt BJ.Plasma D-dimers in the diagnosis of venous thromboembolism,Arch Intern Med,2002:162:747–756.

7. Chen JP,Rowe DW,Ederson BL.Contrasting post-traumatic serial changes for D-dimer and PAI-1 in critically injured patients.Thromb Res,1999,175∼185.

8. Johna S,Cemaj S,O’Callaghan T,Catalano R.Effect of tissue injury on D-dimer levels:a prospective study in trauma patients.Med Sci Monit.2002 Jan;8(1):CR5∼8.

9. Chen Liu, Ying Song, Jingzhong Zhao.Elevated D-dimer and fibrinogen levels in serum of preoperative bone fracture patients.SpringerPlus.2016,5:161–165.

10. Oshima K.J. Usefulness of fibrin clearance products and d-dimer levels as biomarkers that reflect the severity of trauma. Trauma Acute Care Surg,2013,1275∼1278.

11. Sawamura A,Hayakawa M. Disseminated intravascular coagulation with a fibrinolytic phenotype at an early phase of trauma predicts mortality.Thromb Res,2009, 608∼613.

12. Kushimoto S,Gando S.Clinical course and outcome of disseminated intravascular coagulation diagnosed by Japanese Association for Acute Medicine criteria. Comparison between sepsis and trauma. Thromb Haemost.2008, 1099∼1105.

13. Zhang LD,Ma HM.Correlation analysis between plasma D-dimer levels and orthopedic trauma severity. Med J (Engl),2012,3133∼3136.

14. Zhang j. Increased neutrophil elastase, persistent intravascular coagulation and decreased fibrinolytic activity in patients with post-traumatic acute respiratory distress syndrome. Foreign Med Sci·Basic Problems Trauma Surg,1998,1192∼1200.

